# LANTERN: Leveraging Local Ancestry Tracts to Enhance Rare-Variant Aggregate Association Testing

**DOI:** 10.64898/2026.04.24.26351693

**Authors:** Yu Wang, Bjoernar Tuftin, Laura M. Raffield, Bertha Hidalgo, Sarah L. Kerns, Andrew T. DeWan, Suzanne M. Leal, Paul L. Auer

## Abstract

Individuals with admixed ancestry comprise a significant proportion of populations of the Americas. Statistical methods have been developed to specifically leverage local ancestry inference to enhance the power and interpretability of genome-wide association studies in admixed populations. However, no such methods currently exist to test for rare-variant aggregate associations. Here we present LANTERN (Leveraging local ANcestry Tracts to Enhance Rare variaNt aggregate associations), a method that infers the alleles that lie on each ancestral haplotype and conducts rare-variant aggregate association testing in a generalized linear mixed model framework. Through simulation studies we demonstrated that LANTERN achieves proper control of Type 1 error while boosting power to detect associations when causal alleles predominately lie on one ancestral haplotype. Using data from a cohort of African American participants from the Jackson Heart Study, LANTERN identified two genes known to be involved in red-blood cell (RBC) biology when local ancestry information was incorporated. Specifically, a burden of rare alleles on European ancestral haplotypes in *EPO* was associated with both hemoglobin levels (HGB) and RBC counts, whereas a burden of rare alleles on African ancestral haplotypes in *EPB42* was associated with HGB and RBC. In summary, LANTERN (i) allows for the identification of ancestry-specific rare-variant associations; and (ii) enhances rare-variant association signals compared to an analysis that ignores local ancestry. LANTERN is implemented in R and is freely available on GitHub.

## Main Text

Individuals with admixed genetic ancestry comprise a significant proportion of populations of the Americas including the United States.^1^ Additional countries with high proportion of admixed individuals outside the Americas include, but are not limited to, South Africa and the Philippines. However, individuals with admixed ancestry represent a disproportionally small fraction of individuals included in genome-wide association studies (GWAS).^2^ Recent efforts, such as the NHLBI Trans Omics for Precision Medicine (TOPMed) and the NIH All of Us Research programs, have made strides in prioritizing the inclusion of admixed individuals in large-scale genetic studies.^3,4^ Along with these efforts in data collection, investigators have developed new statistical methods for leveraging local ancestry inference to inform genetic association studies.^5^ One such method, “Tractor”, analyzes admixed genomes to detect phenotype associations in a regression framework by (1) including inferred local ancestries as covariates; and (2) counting the number of risk alleles on each inferred ancestral haplotype. This method has proven powerful in identifying ancestry-specific effects for individual variants and improving resolution of association signals.

Tractor relies on two key pieces of information: (1) inferred local ancestry at every marker being tested; and (2) phased genotypes, i.e., haplotype resolved genotypes, so that alleles can be confidently placed on ancestral haplotypes. It is well known that when parental data is unavailable and the minor allele frequency (MAF) of a variant decreases, the confidence of statistical phasing diminishes.^6^ For variants that are only observed in a single individual (i.e., singletons), there are no template haplotypes that contain the rare allele and thus the placement of the rare allele on one ancestral haplotype is completely random. Therefore, association methods that rely on statistical phasing are inaccurate for rare-variants. Additionally, rare-variant association studies rarely test one marker for an association at a time, as is done in standard GWAS but rather aggregate rare-variants into a unit for analysis. Often individual genes are the unit of analysis or genes and their promoters and/enhancers. Typically, rare-variant aggregate association tests only include a subset of variants within the unit of analysis, such as nonsynonymous, frameshift, splice-site, or predicted loss-of-function variants. Aggregate rare-variant association tests include both a “burden” test where one sums the number of rare alleles to create a burden score that is tested for association with a phenotype or a “variance component” test where one tests for the heterogeneity of effect of rare-variants on the phenotype. A large number of aggregate rare-variant association tests have been developed, each with their own advantages and disadvantages,^7^ but none that leverage local ancestry tracts in admixed individuals. Here, we present a new method LANTERN (Leveraging local ANcestry Tracts to Enhance Rare variaNt aggregate associations), that infers the alleles that occupy each ancestral haplotype and conducts rare-variant aggregate association testing in a linear mixed model (LMM) or generalized LMM framework.^8^

For simplicity, consider a set of individuals with two-way African (AF) and European (EU) admixture. Like other rare-variant tests, we use a MAF cut-off (e.g., MAF<0.01) where only variants meeting that cutoff and of specific functional class(es) (e.g. predicted loss of function) variants are analyzed. We construct two models, one for each ancestry group, that are each similar in form to the Tractor model:

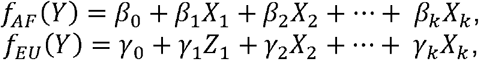

where *f*_*AF*_ (*Y*) and *f*_*EU*_ (*Y*) represent some function of the outcome variable Y (e.g., logit(Y)), *X*_1_ represents the number of copies of the rare allele that are present on the AF haplotype, *Z*_1_ the number of copies of the rare allele that are present on the EU haplotype, and *X*_2_, …, *X*_*k*_ represent covariates such as age, sex, global ancestry estimates, or local ancestry estimates.

We extend this model in two ways: (1) include a random effect term *g*, to *f*_*AF*_ (Y) and *f*_*EU*_ (*Y*), respectively, where *g*∼*N*(0, 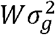), *W* is an empirical kinship matrix, and 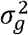 is the corresponding variance component parameter; and (2) assign rare alleles to ancestral haplotypes as shown in **Table 1**. Then both null models (i.e., *β*_1_ = 0, and *γ*_1_ = 0) are fit using restricted maximum likelihood (REML) and score statistics are constructed using the variant-set mixed model association test (SMMAT) machinery.^8^

**Table 1.** Determining the probabilities that the rare allele lies on an African (AF) or European (EU) ancestral haplotype for an individual that has 2-way AF and EU admixture.

| Local Ancestry | Genotype | # AF Alleles | #EU Alleles | Number of observations |
| --- | --- | --- | --- | --- |
| AF/AF | AA | 2 | 0 | $N_1$ |
| | AB | 1 | 0 | $N_2$ |
| | BB | 0 | 0 | $N_3$ |
| AF/EU | AA | 1 | 1 | $N_4$ |
| | AB | $P_1$ | $P_2$ | $N_5$ |
| | BB | 0 | 0 | $N_6$ |
| EU/EU | AA | 0 | 2 | $N_7$ |
| | AB | 0 | 1 | $N_8$ |
| | BB | 0 | 0 | $N_9$ |
| A=rare allele; B=common allele; N= number of samples with the specified local ancestry and genotype. |  |  |  |  |
| $P_1 = \frac{\text{\# times A is on the AF haplotype}}{\text{\# times A is observed}} = \frac{2N_1 + N_2 + N_4}{2N_1 + N_2 + 2N_4 + 2N_7 + N_8}$ | | | | |
| $P_2 = \frac{\text{\# times A is on the EU haplotype}}{\text{\# times A is observed}} = \frac{N_4 + 2N_7 + N_8}{2N_1 + N_2 + 2N_4 + 2N_7 + N_8}$ | | | | |
| $1 = P_1 + P_2$ | | | | |

This framework permits real-value quantities for *X*_1_ and *Z*_1_, which is necessary when *P*_1_ and *P*_2_ are estimated (see caption to **Table 1**). Note that there are situations when we will not have additional information with which to estimate *P*_1_ and *P*_2_. For instance, if a variant is observed with *N*_5_=1 and all other values in **Table 1** equal to zero. In other words, a singleton at a locus with AF/EU inferred local ancestry. In this case we assume that the A allele has a 50% chance of occurring on the AF haplotype and a 50% chance of occurring on the EU haplotype (i.e., that 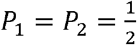), though our implementation of LANTERN allows for user-defined values of this weighting (if, for instance, the user prefers to input the global ancestry estimates for *P*_1_and *P*_2_).

Thus defined, LANTERN fits seamlessly into rare-variant burden and variance component testing as implemented in the Set Mixed Model Association Tests (SMMAT) model. Specifically, SMMAT constructs score statistics *U*_*k*_ for every variant *k* that is included in the aggregate test for a gene/region. *U*_*k*_ is used to evaluate the null hypothesis that the effect of the *k*^*th*^ variant is non-zero and the *U*_*k*_ statistics are combined across all variants within a gene/region to construct burden, variance components, and combined (i.e., “optimal”) tests. With the allocation of alleles in LANTERN as described above, SMMAT is applied twice: once for the rare alleles on the AF haplotypes and once for the rare alleles on the EU haplotypes, resulting in 2 score statistics for each variant *k* (*U*_1*k*_, *U*_2*k*_) that are used to evaluate the two null hypotheses *H*_01_: *β*_1_ = 0, *H*_02_ : *γ*_1_ = 0, respectively. This amounts to two tests of ancestry specific effects, one each for AF and EU. Then we use the SMMAT machinery to combine score statistics to create burden, variance component, and optimal tests for the gene/region being analyzed. To combine p-values across ancestries, the Cauchy combination method is used,^9^ weighting the p-values by the total number of ancestral haplotypes observed in the gene being analyzed. User defined weights are also permitted.

To evaluate the performance of LANTERN, we performed comprehensive simulations using genetic data from the Jackson Heart Study (JHS),^10^ a cohort of 5,306 admixed African Americans living in and near Jackson, Mississippi. Sequencing of 3,406 participants in the JHS cohort was performed as part of the TOPMed program (see **Web Resources**)^4^ local ancestry was estimated via RFmix,^11^ as previously described,^12^ and the empirical kinship matrix was estimated using KING identity by descent (IBD) segment inference.^13^ Note that the JHS cohort includes both closely and distantly related individuals. We filtered the whole genome sequence (WGS) data to variants categorized as “missense”, “nonsense” or “splice” in VEP^14^ v100 predicted consequence for ENSEMBL transcripts, in the WGSA^15^ TOPMed annotation files. To demonstrate the usefulness of LANTERN, we used LANTERN to investigate red-blood cell count (RBC) and hemoglobin concentration (HGB), two heritable quantitative traits with well-documented genetic loci that differ across ancestries. To do so, we filtered the JHS samples to those that had phenotypic data on RBC and HGB. In the simulations, LANTERN was benchmarked against ignoring local ancestry estimates and simply running SMMAT on all observed variants. For all analyses (both in the power and Type 1 error simulations and the real-world data analysis) we only included variants with an observed MAF<1%. The Institutional Review Boards at Jackson State University, Tougaloo College, and the University of Mississippi Medical Center approved the original study protocol, and all participants provided written informed consent. Ongoing IRB review for study activities is through the University of Mississippi Medical Center and Wake Forest University Health Sciences.

To evaluate the performance of LANTERN, we considered a random selection of nine genes that have ≥ 10 rare-variants on chromosome 15 in the JHS data (see **Table S1**). This region was chosen due to its relative lack of large-scale linkage disequilibrium blocks (LD) in both AF and EU populations. Analysis was limited to nine genes to keep computational expense feasible.

Using these nine genes, Type I error and power analyses were conducted as follows: let *n* denote the sample size, *v*_*s*_ represent the set of observed variants within gene *s*, and **x**_*obs*_ represent the observed genotype matrix for gene *s*. Using the rules defined in **Table 1**, we construct ancestry specific genotype matrices **x**_*AF*_ and **x**_*EU*_. Let *κ* denote the kinship matrix between all samples. The vector of effect sizes for variants within the gene *s* is defined as 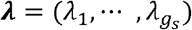, where the variance of the effect size distribution is *σ*_*γ*_ . We include an indicator variable representing the causality of each variant (i.e., 1=causal, 0=non-causal) defined as 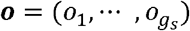, with the probability of a variant being causal = *p*. The identity matrix is denoted by *I*. Variance components include the kinship variance component 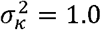 and the noise variance component 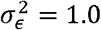. In our simulations, we set *P*_1_ =*P*_2_ =0 (from **Table 1**). Effect sizes within each gene were generated independently from the absolute value of a normal distribution, 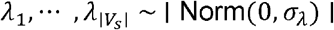, implying an average effect size of 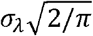. Variant causality indicators were generated independently as 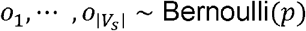. Continuous phenotypes were then simulated from a multivariate normal distribution, 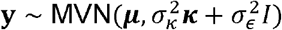, where the mean vector is defined as ***μ* =** (***λ*** ∘ ***o***)**x**, with different causal configurations considered. We considered scenarios where only alleles on AF haplotypes have causal effects: ***μ***_*AF*_ = (***λ***_*AF*_ ∘ ***o***_*AF*_)**x**_*AF*_; only alleles on EU haplotypes have causal effects: ***μ***_*EU*_ = (***λ***_*EU*_ ∘ ***o***_*EU*_)**x**_*EU*_; and alleles have causal effects regardless of which ancestral haplotypes they occupy: ***μ***_*Mixed*_ = (***λ***_*Mixed*_ ∘ ***o***_*Mixed*_)**x**_*obs*_ . In the first two instances (***μ***_*AF*_, and ***μ***_*EU*_), we did not include certain singletons (*N*_5_ in **Table 1**) in the simulations to ensure that causal effects were unambiguously assignable to one ancestry.

Following phenotype simulation, SMMAT models were fitted and adjusted p-values were obtained by testing associations using AF genotypes (**y**∼ **x**_*AF*_ → pvalue_AF_), EU ancestry genotypes (**y**∼ **x**_*EU*_ → pvalue_EU_), and combined observed genotypes (**y**∼ **x**_*obs*_ → pvalue_obs_). These p-values represent the SKAT-O tests from SMMAT. In evaluating Type I error, we simply set ***o*** = 0, use the real observed variant matrix **x**_*obs*_ and the kinship matrix ***κ*** from the JHS cohort, and ran 1 million simulations to estimates of Type 1 error at the 5×10^-6^ level, for each of the 9 genes we considered. We randomly down-sampled to 1000 subjects due to the large computation burden of running 1 million simulations for the Type 1 error estimation.

As shown in **Table 2**, the AF LANTERN model (LANTERN-AF), the EU LANTERN model (LANTERN-EU), and the Cauchy-Weighted LANTERN model (LANTERN-Cauchy) all obtained the approximately nominal Type 1 error level, with performance very similar to that of SMMAT (i.e., ignoring local ancestry altogether). Quantile-quantile plots were generated using 1 million simulations, for each gene (**Supplementary Figures S1-S4)**.

**Table 2.** Gene-level Type I error across methods with a significance level of 5×10^-6^. The last row reports the mean and standard deviation (SD) across genes for each method.

| Gene | LANTERN-AF | LANTERN-EU | LANTERN-Cauchy | SMMAT |
| --- | --- | --- | --- | --- |
| *ANKRD63* | $4.00 \times 10^{-6}$ | $7.00 \times 10^{-6}$ | $4.00 \times 10^{-6}$ | $2.00 \times 10^{-6}$ |
| *FOXB1* | $8.00 \times 10^{-6}$ | $6.00 \times 10^{-6}$ | $8.00 \times 10^{-6}$ | $7.00 \times 10^{-6}$ |
| *GOLGA6L9* | $7.00 \times 10^{-6}$ | $4.00 \times 10^{-6}$ | $7.00 \times 10^{-6}$ | $8.00 \times 10^{-6}$ |
| *ISL2* | $4.00 \times 10^{-6}$ | $3.00 \times 10^{-6}$ | $5.00 \times 10^{-6}$ | $3.00 \times 10^{-6}$ |
| *LDHAL6B* | $3.00 \times 10^{-6}$ | $3.00 \times 10^{-6}$ | $3.00 \times 10^{-6}$ | $4.00 \times 10^{-6}$ |
| *ODF3L1* | $2.00 \times 10^{-6}$ | 0.00 | $1.00 \times 10^{-6}$ | $2.00 \times 10^{-6}$ |
| *OR4F6* | $2.00 \times 10^{-6}$ | $5.00 \times 10^{-6}$ | $2.00 \times 10^{-6}$ | $3.00 \times 10^{-6}$ |
| *RAMAC* | $3.00 \times 10^{-6}$ | $4.00 \times 10^{-6}$ | $3.00 \times 10^{-6}$ | $8.00 \times 10^{-6}$ |
| *SPESP1* | $4.00 \times 10^{-6}$ | $7.00 \times 10^{-6}$ | $5.00 \times 10^{-6}$ | $3.00 \times 10^{-6}$ |
| Mean (SD) | $4.11 \times 10^{-6}$<br>($2.09 \times 10^{-6}$) | $4.33 \times 10^{-6}$<br>($2.24 \times 10^{-6}$) | $4.22 \times 10^{-6}$<br>($2.28 \times 10^{-6}$) | $4.44 \times 10^{-6}$<br>($2.51 \times 10^{-6}$) |

To evaluate the statistical power of LANTERN, we considered the same set of nine genes on chromosome 15 and generated phenotypes using the fully observed **x**_obs_ and ***κ*** from the study cohort, setting the causality rate *p* = (0.25, 0.50, 0.75), and *σ*_*λ*_ such that the average effect size was (1.0, 1.5, 2.0, 2.5, 3.0).

Three types of power simulations were carried out: (1) only alleles on AF haplotypes exerted phenotypic effects; (2) only alleles on EU haplotypes exerted phenotypic effects; and (3) alleles exerted phenotypic effects regardless of the ancestral haplotype on which they were located. **Figure 1** illustrates the differences in power for each method using *ANKRD63* as an example. When only alleles on AF haplotypes exerted phenotypic effects, the LANTERN-Cauchy and LANTERN-AF models demonstrated the highest power, followed by SMMAT. Correctly, the power of LANTERN-EU was equal to the Type 1 error rate. This pattern held up regardless of the proportion of causal variants. When only alleles on EU haplotypes exerted phenotypic effects, LANTERN-EU had by far the highest power, followed by SMMAT and LANTERN-Cauchy. LANTERN-AF had power equal to the Type 1 error rate in this situation. This result points to a particular advantage of the general LANTERN framework. When one ancestry is consistently under-represented in a sample of admixed genomes, that ancestry-specific LANTERN test will provide a large boost in power compared to ignoring ancestry altogether. In this scenario, ignoring ancestry effectively allows the ancestry group that dominates the admixture proportions to wash out the ancestry-specific effects of the non-dominant ancestry group. When local ancestry did not influence the generation of phenotypes, LANTERN-Cauchy and SMMAT performed similarly with respect to power. Similar patterns were observed for the simulations performed using the other eight genes (**Supplementary Figures 5-7**).

**Figure 1.**
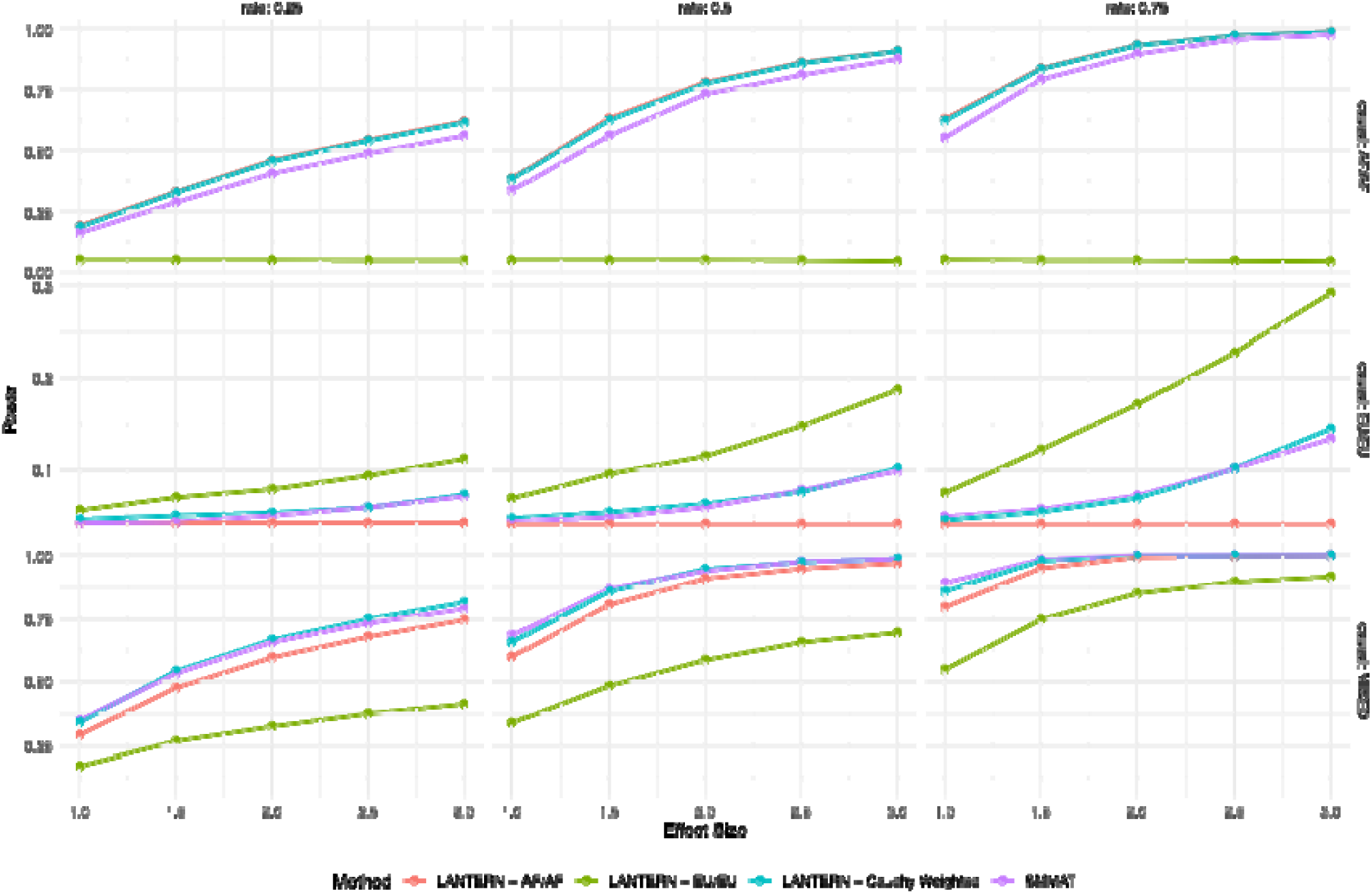
Power to detect rare-variant associations in the *ANKRD63* gene. The results for LANTERN-AF are shown in red, LANTERN-EU in green, LANTERN-Cauchy in blue, and SMMAT in purple. Causal variants were simulated in 25% of variants (left panel), 50% of variants (middle panel), and 75% of variants (right panel). Causal alleles were placed exclusively on AF haplotypes in the top panel, exclusively on EU haplotypes middle panel, and regardless of ancestral haplotypes (bottom panel). Simulated effect sizes are shown on the horizontal axis.

To demonstrate the utility of LANTERN, we analyzed hemoglobin (HGB) concentrations and total RBC counts in 3,314 participants with measured phenotypes in JHS. HGB (N=3,312) was defined as the mass per volume (grams per deciliter) of hemoglobin in the blood and RBC count (N=2972) was defined as the count of RBCs in the blood, by number concentration in millions per microliter.

Though our sample of 3,314 African Americans was under-powered to detect novel rare-variant associations, i.e., exome-wide significant 2.71×10^-6^ (a Bonferroni correction for testing 18,464 genes) with HGB or RBC, the p-values from LANTERN were not systematically inflated or deflated (see **Supplementary Figures 8-9**). Additionally, the results for two genes known to be involved in red blood-cell biology [*EPBA42* (Protein 4.2, Erythrocytic; OMIM 177070) and *EPO* (Erythropoietin; OMIM 133170)] highlight the advantages of LANTERN compared to conducting rare-variant association testing without consideration of local ancestry.

*EPO* encodes a glycoprotein hormone that regulates RBC production and HGB concentration. Prior studies have identified rare, missense variants in *EPO* as genetic determinants of red blood cell traits.^16^ In our analysis of the JHS data, there were 9 rare-variants included in the LANTERN test for *EPO* (see **Table S2**). In **Table 3**, we see that the associations with both HGB and RBC in JHS were limited to alleles on EU haplotypes. In particular, EU haplotype-specific alleles show a strong association with RBC. The LANTERN-EU tests were the only method that returned nominally significant p-values. The associations on AF haplotypes were null, as were the associations ignoring ancestry. In this case, LANTERN provides a significant boost in power compared to either ignoring ancestry or testing only AF-specific signals.

**Table 3.** Aggregate rare-variant association p-values for *EPB42* and *EPO* with HGB and RBC traits.

| Gene | Trait | LANTERN-AF<br>Burden | LANTERN-AF<br>SKAT-O | LANTERN_EU<br>Burden | LANTERN-EU<br>SKAT-O | SMMAT<br>Burden | SMMAT<br>SKAT-O | Cauchy<br>SKAT-O |
| --- | --- | --- | --- | --- | --- | --- | --- | --- |
| *EPB42* | HGB | 0.00057 | 0.0043 | 0.9015 | 0.9380 | 0.0011 | 0.0077 | 0.0006 |
|  | RBC | 0.00151 | 0.0078 | 0.7152 | 0.9121 | 0.0044 | 0.0211 | 0.0016 |
| *EPO* | HGB | 0.5448 | 0.5324 | 0.0976 | 0.0760 | 0.6529 | 0.4255 | 0.4970 |
|  | RBC | 0.9165 | 0.5508 | 0.0097 | 0.01994 | 0.1823 | 0.10708 | 0.8551 |

*EPB42* is a protein that interacts with the RBC membrane cytoskeleton.^17^ Rare variants in *EPB42* are known to be associated with reduced hemoglobin and Spherocytosis Type 5 (OMIM 612690), a phenotype characterized by hemolytic anemia and abnormally shaped, fragile red blood cells. There were 41 variants included in the LANTERN association test for *EPB42* (see Table **S3**). As shown in **Table 3**, the associations with both HGB and RBC in JHS were limited to alleles on AF haplotypes. In this scenario, the LANTERN-AF and Cauchy tests returned nominally significant p-values that were smaller than testing for association with SMMAT which ignores ancestry. The associations for alleles on EU haplotypes appeared null. Here, we see that LANTERN provides a power boost when testing for the association with alleles on AF haplotypes, even when on average the ancestry of the sample is roughly 80% African.

In addition to scenarios with two-way ancestry, LANTERN is extensible to multi-way admixture; our current implementation extends up to 5-way admixture. As can be seen in both our simulations and real-data analysis, the power gains with LANTERN are most realized in identifying ancestry-specific effects in ancestries that contribute small amounts of admixture in a particular sample of individuals. For instance, the Hispanic Community Health Study/Study of Latinos (HCHS/SOL) is a population-based cohort of Hispanic/Latinos ascertained from the Bronx, New York; Chicago, Illinois; Miami, Florida, and San Diego, California.^18^ Here, the term “Hispanic/Latinos” is used as a population descriptor and captures a diverse background distribution, with major groups being Mexican, Cuban, Puerto Rican, Dominican, Central American, and South American. Previous studies have shown that Amerindian ancestry for the HCHS/SOL study subjects ranges from <5% to ∼80% with an average of 30.5%.^19^ For a study like HCHS/SOL, LANTERN may be particularly effective at identifying an aggregate of rare-alleles specific to Amerindian haplotypes that are associated with a trait of interest.

Because of its ability to detect ancestry-specific rare-variant associations, the development of LANTERN may also assist with portability of polygenic risk scores (PRS). Prior work has shown that the inclusion of rare-variants may boost the predictive ability of PRS^20^ and that transferability is improved when PRS include variants that are discovered from diverse populations.^21^

Some of the most impactful instances of human genetic studies leading to novel drug discovery have been made from population-specific rare-variant associations. For instance, the discovery of rare variants in African Americans in the *PCKS9* gene^22^ that were associated with low-density lipoprotein (LDL) cholesterol levels directly led to the development of monoclonal antibodies to treat individuals with high LDL-levels.^23^ Rare-variants observed in African Americans in the *APOC3* gene^24,25^ were also associated with triglyceride levels and risk for cardiovascular disease and led to the development of new therapies.^26^ Thus, implementation of methods such as Tractor and LANTERN holds the promise for more discoveries that could lead to novel therapeutics.

As with the Tractor method for common variants, LANTERN implicitly accounts for local gene-gene interaction effects by essentially stratifying the association test by ancestry tracks. These ancestry tracks are often quite large and extend across multiple genes. LANTERN can also be used with the output from statistical phasing algorithms as well, if investigators prefer to use those assignments of alleles to ancestral haplotypes rather than the rules dictated in **Table 1**. Finally, we note that LANTERN can be used to analyze case-control phenotypes just as easily as with quantitative traits.

In summary, LANTERN is an approach for testing aggregate rare-variant associations that leverages admixture to identify genetic effects that are specific to an ancestral haplotype. As large-scale sequencing studies continue to generate WGS data on diverse sets of individuals, many of which are characterized by admixed ancestry, similar statistical methods will be needed to fully capture the richness of genotype-phenotype associations across populations.

## Data Availability

Data from the JHS TOPMed project is available on dbGaP under accession phs000964.

## Data and code availability

LANTERN is freely available to download from source via GitHub (see **Web Resources**). Full installation and execution instructions, as well as example input data, are on GitHub. Data from the JHS TOPMed project is available on dbGaP under accession phs000964.

## Acknowledgements

This work was supported by NIH grant R01DC017712 (S.M.L, A.T.D., P.L.A.) LMR was supported by R01HL146500. The Jackson Heart Study is supported and conducted in collaboration with Tougaloo College (75N92025D00038), Jackson State University (75N92025D00039), University of Southern Mississippi (75N92025D00040), G.A. Carmichael Family Health Center (75N92025D00041), Wake Forest University Health Sciences (75N92025D00036), and the University of Mississippi Medical Center (75N92025D00037) contracts from the National Heart Lung and Blood Institute (NHLBI) with additional support from the National Institute of Minority Health and Health Disparities (NIMHD).Molecular data for the Trans-Omics in Precision Medicine (TOPMed) program was supported by the National Heart, Lung and Blood Institute (NHLBI). Genome sequencing for “NHLBI TOPMed: The Jackson Heart Study” (phs000964.v1.p1) was performed at the Northwest Genomics Center (HHSN268201100037C). Core support including centralized genomic read mapping and genotype calling, along with variant quality metrics and filtering were provided by the TOPMed Informatics Research Center (3R01HL-117626-02S1; contract HHSN268201800002I). Core support including phenotype harmonization, data management, sample-identity QC, and general program coordination were provided by the TOPMed Data Coordinating Center (R01HL-120393; U01HL-120393; contract HHSN268201800001I). We gratefully acknowledge the studies and participants who provided biological samples and data for TOPMed. The authors thank the participants and staff of the Jackson Heart Study for their important contributions.

## Author Contributions

P.L.A, A.T.D., and S.M.L. designed the study and oversaw the simulations and data analysis. Y.W. wrote the code and ran the simulations. B.T. ran the data analysis in JHS supervised by L.M.R. P.L.A. and Y.W. wrote the manuscript. All authors provided critical review and revisions of the manuscript.

## Declaration of Interests

The authors declare no competing interests.

## Web Resources

SMMAT (https://www.rdocumentation.org/packages/GMMAT/versions/1.4.2/topics/SMMAT)

LANTERN (https://github.com/yuw444/lantern)

TOPMed Methods (https://topmed.nhlbi.nih.gov/topmed-whole-genome-sequencing-methods-freeze-8)

## Notes

### Competing Interest Statement

The authors have declared no competing interest.

### Author Declarations

Data from the Jackson Heart Study TOPMed project is available on dbGaP under accession phs000964.

## References

1. Bryc, K., Durand, E. Y., Macpherson, J. M., Reich, D. & Mountain, J. L. The genetic ancestry of African Americans, Latinos, and European Americans across the United States. Am. J. Hum. Genet. 96, 37–53 (2015).

2. Sirugo, G., Williams, S. M. & Tishkoff, S. A. The Missing Diversity in Human Genetic Studies. Cell 177, 26–31 (2019).

3. All of Us Research Program Genomics Investigators. Genomic data in the All of Us Research Program. Nature 627, 340–346 (2024).

4. Taliun, D. et al. Sequencing of 53,831 diverse genomes from the NHLBI TOPMed Program. Nature 590, 290–299 (2021).

5. Atkinson, E. G. et al. Tractor uses local ancestry to enable the inclusion of admixed individuals in GWAS and to boost power. Nat. Genet. 53, 195–204 (2021).

6. Hofmeister, R. J., Ribeiro, D. M., Rubinacci, S. & Delaneau, O. Accurate rare variant phasing of whole-genome and whole-exome sequencing data in the UK Biobank. Nat. Genet. 55, 1243–1249 (2023).

7. Lee, S., Abecasis, G. R., Boehnke, M. & Lin, X. Rare-variant association analysis: study designs and statistical tests. Am. J. Hum. Genet. 95, 5–23 (2014).

8. Chen, H. et al. Efficient Variant Set Mixed Model Association Tests for Continuous and Binary Traits in Large-Scale Whole-Genome Sequencing Studies. Am. J. Hum. Genet. 104, 260–274 (2019).

9. Liu, Y. & Xie, J. Cauchy combination test: a powerful test with analytic p-value calculation under arbitrary dependency structures. J. Am. Stat. Assoc. 115, 393–402 (2020).

10. Sempos, C. T., Bild, D. E. & Manolio, T. A. Overview of the Jackson Heart Study: a study of cardiovascular diseases in African American men and women. Am. J. Med. Sci. 317, 142–146 (1999).

11. Maples, B. K., Gravel, S., Kenny, E. E. & Bustamante, C. D. RFMix: A Discriminative Modeling Approach for Rapid and Robust Local-Ancestry Inference. Am. J. Hum. Genet. 93, 278–288 (2013).

12. Cruz, D. E. et al. Admixture-mapping analysis reveals genetic determinants of the human plasma proteome. HGG Adv. 7, 100529 (2026).

13. Manichaikul, A. et al. Robust relationship inference in genome-wide association studies. Bioinformatics 26, 2867–2873 (2010).

14. McLaren, W. et al. The Ensembl Variant Effect Predictor. Genome Biol. 17, 122 (2016).

15. Liu, X. et al. WGSA: an annotation pipeline for human genome sequencing studies. J. Med. Genet. 53, 111–112 (2016).

16. Auer, P. L. et al. Rare and low-frequency coding variants in CXCR2 and other genes are associated with hematological traits. Nat. Genet. 46, 629–634 (2014).

17. Azim, A. C. et al. Human erythrocyte dematin and protein 4.2 (pallidin) are ATP binding proteins. Biochemistry 35, 3001–3006 (1996).

18. Lavange, L. M. et al. Sample design and cohort selection in the Hispanic Community Health Study/Study of Latinos. Ann. Epidemiol. 20, 642–649 (2010).

19. Browning, S. R. et al. Local Ancestry Inference in a Large US-Based Hispanic/Latino Study: Hispanic Community Health Study/Study of Latinos (HCHS/SOL). G3 6, 1525–1534 (2016).

20. Dornbos, P. et al. A combined polygenic score of 21,293 rare and 22 common variants improves diabetes diagnosis based on hemoglobin A1C levels. Nat. Genet. 54, 1609–1614 (2022).

21. Cavazos, T. B. & Witte, J. S. Inclusion of variants discovered from diverse populations improves polygenic risk score transferability. HGG Adv. 2, 100017 (2021).

22. Cohen, J. C., Boerwinkle, E., Mosley, T. H. & Hobbs, H. H. Sequence variations in PCSK9, low LDL, and protection against coronary heart disease. N. Engl. J. Med. 354, 1264–1272 (2006).

23. Ballantyne, C. M. et al. Efficacy and Safety of Oral PCSK9 Inhibitor Enlicitide in Adults With Heterozygous Familial Hypercholesterolemia: A Randomized Clinical Trial. JAMA 335, 129–139 (2026).

24. TG and HDL Working Group of the Exome Sequencing Project, National Heart, Lung, and Blood Institute et al. Loss-of-function mutations in APOC3, triglycerides, and coronary disease. N. Engl. J. Med. 371, 22–31 (2014).

25. Auer, P. L. et al. Guidelines for Large-Scale Sequence-Based Complex Trait Association Studies: Lessons Learned from the NHLBI Exome Sequencing Project. Am. J. Hum. Genet. 99, 791–801 (2016).

26. Lightbourne, M. et al. Volanesorsen, an antisense oligonucleotide to apolipoprotein C-III, increases lipoprotein lipase activity and lowers triglycerides in partial lipodystrophy. J. Clin. Lipidol. 16, 850–862 (2022).

