## Supplementary Figures for "LANTERN: Leveraging Local Ancestry Tracts to Enhance Rare-Variant Aggregate Association Testing"

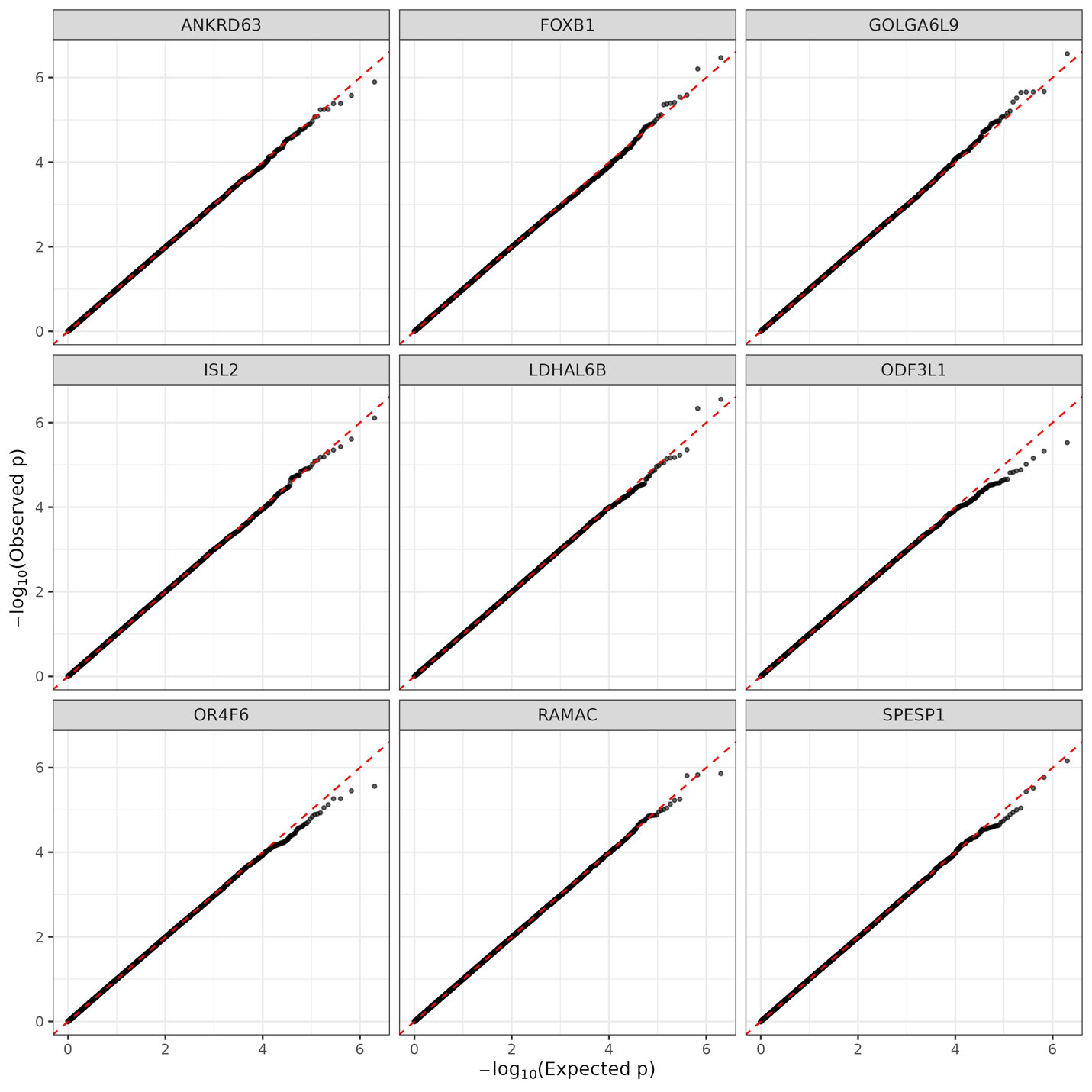

**Figure S1**: QQ-plots for the Type 1 error simulations across all 9 genes on chromosome 15, for the LANTERN-AF/AF model.

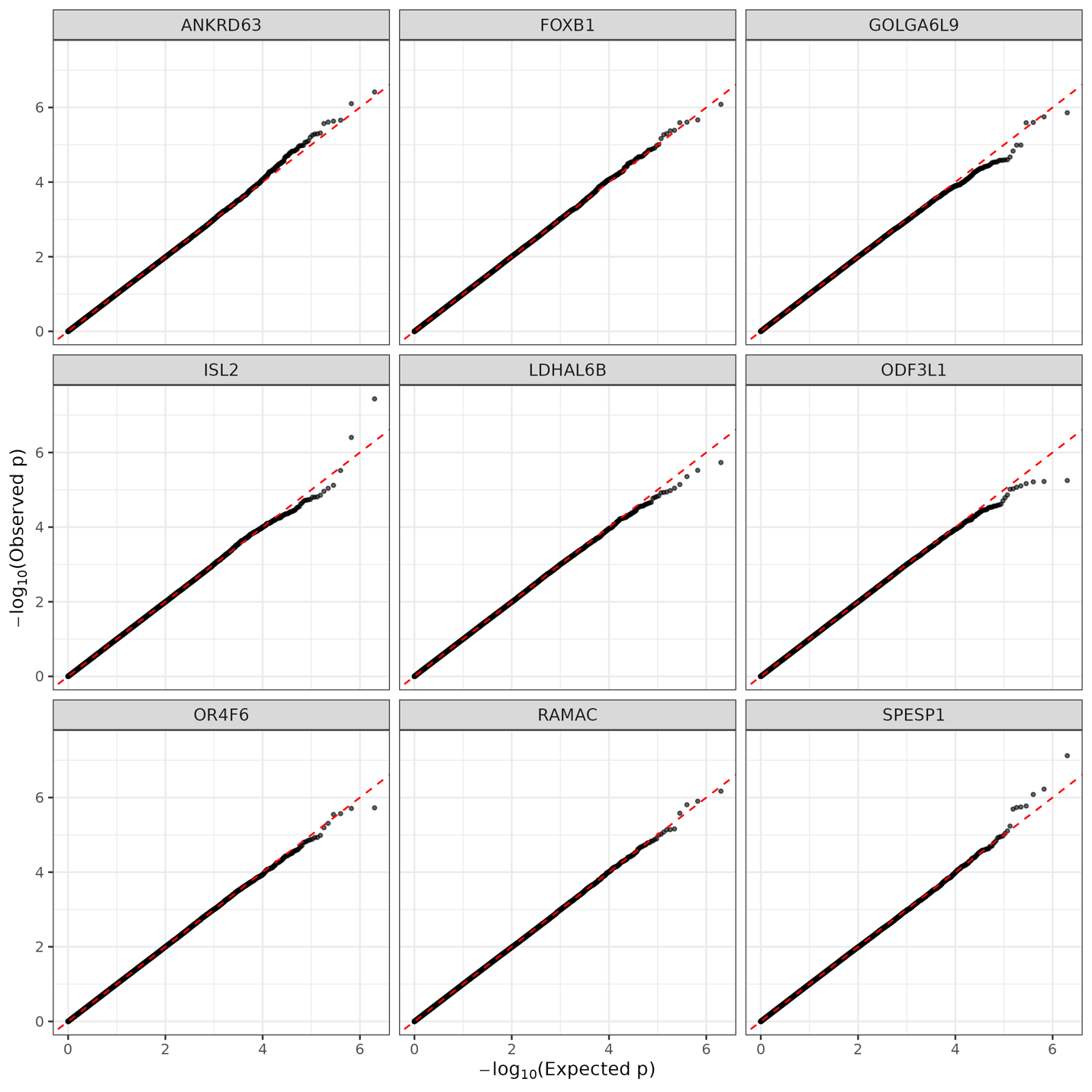

**Figure S2**: QQ-plots for the Type 1 error simulations across all 9 genes on chromosome 15, for the LANTERN-EU/EU model.

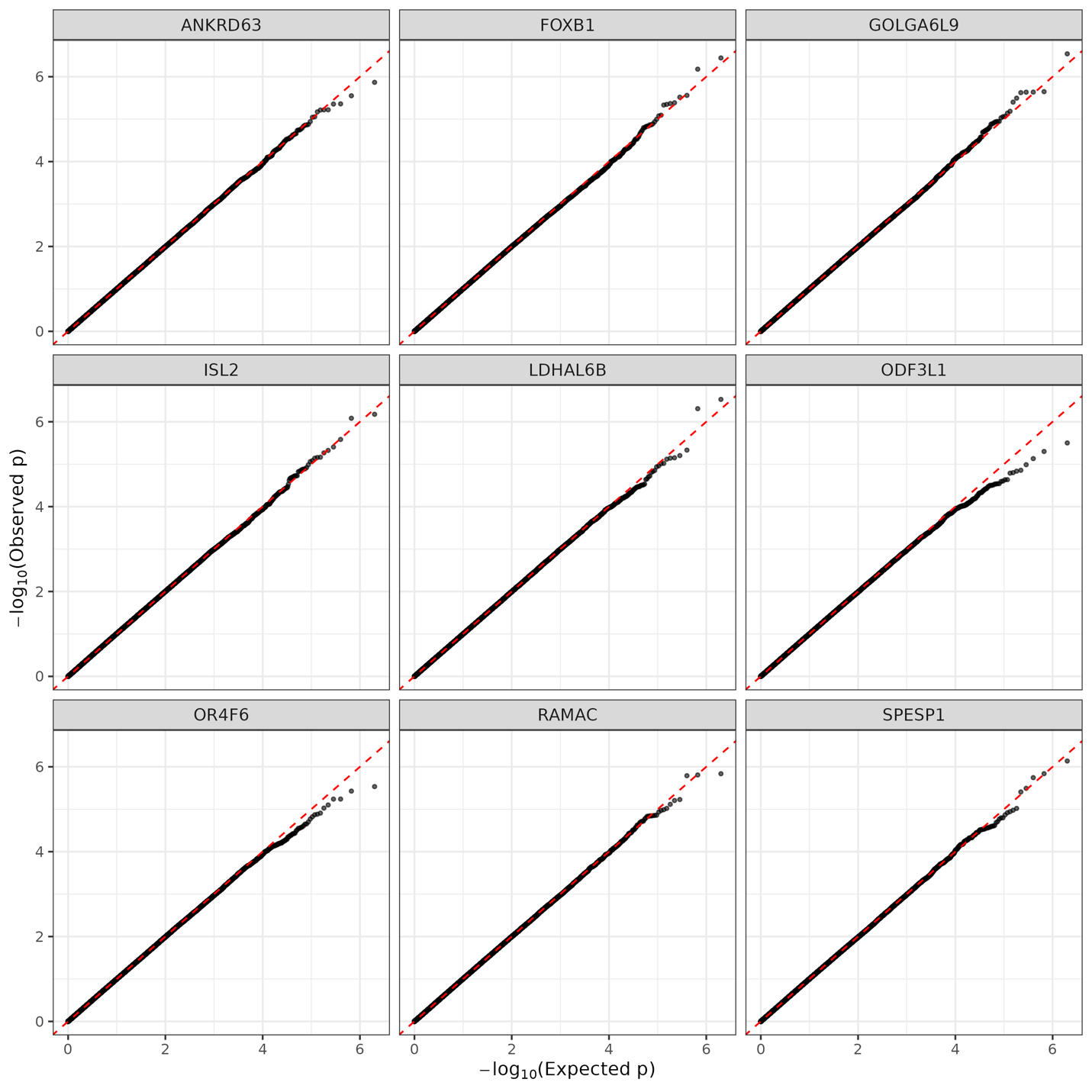

**Figure S3**: QQ-plots for the Type 1 error simulations across all 9 genes on chromosome 15, for the LANTERN-Cauchy model.

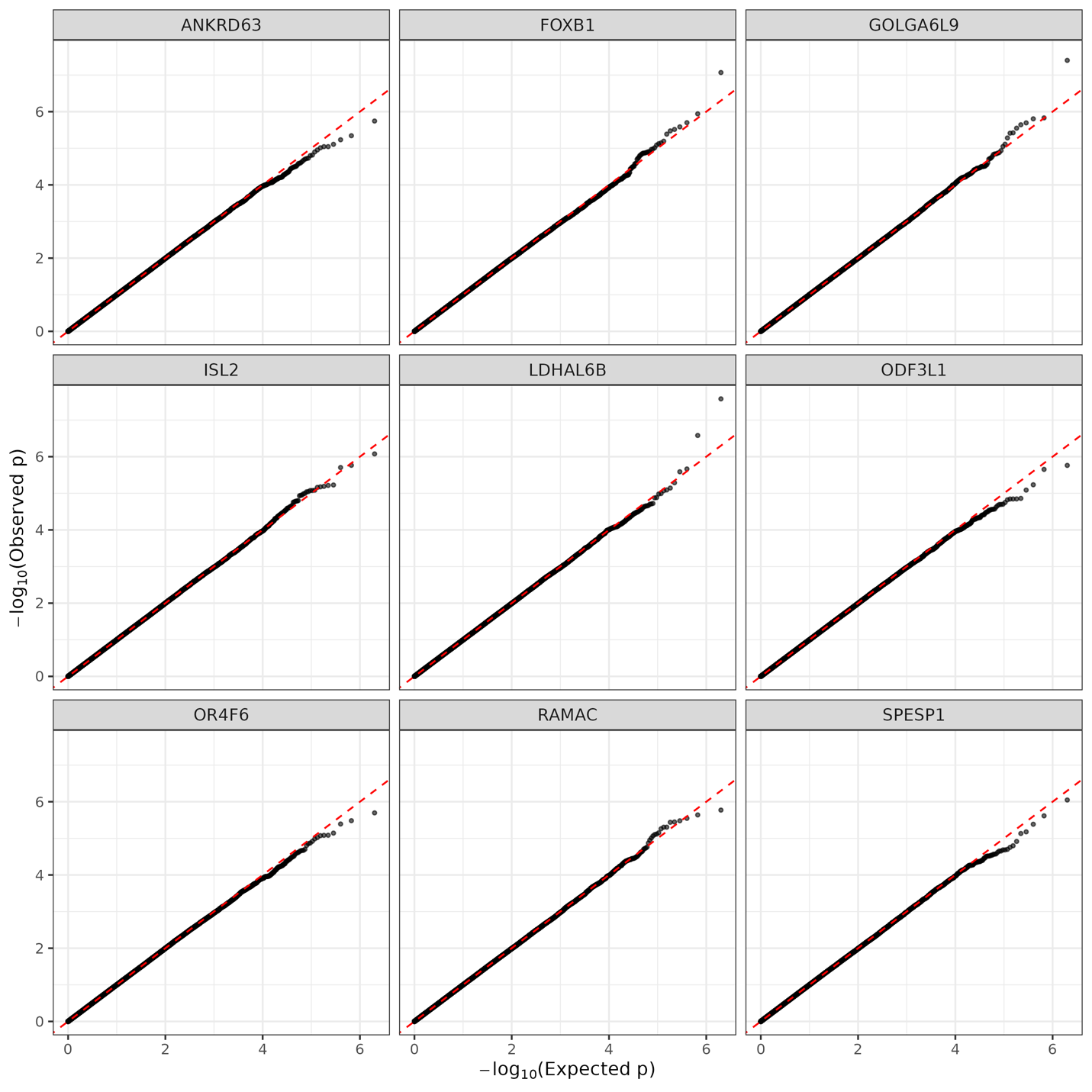
 **Figure S4**: QQ-plots for the Type 1 error simulations across all 9 genes on chromosome 15, for the SMMAT model.

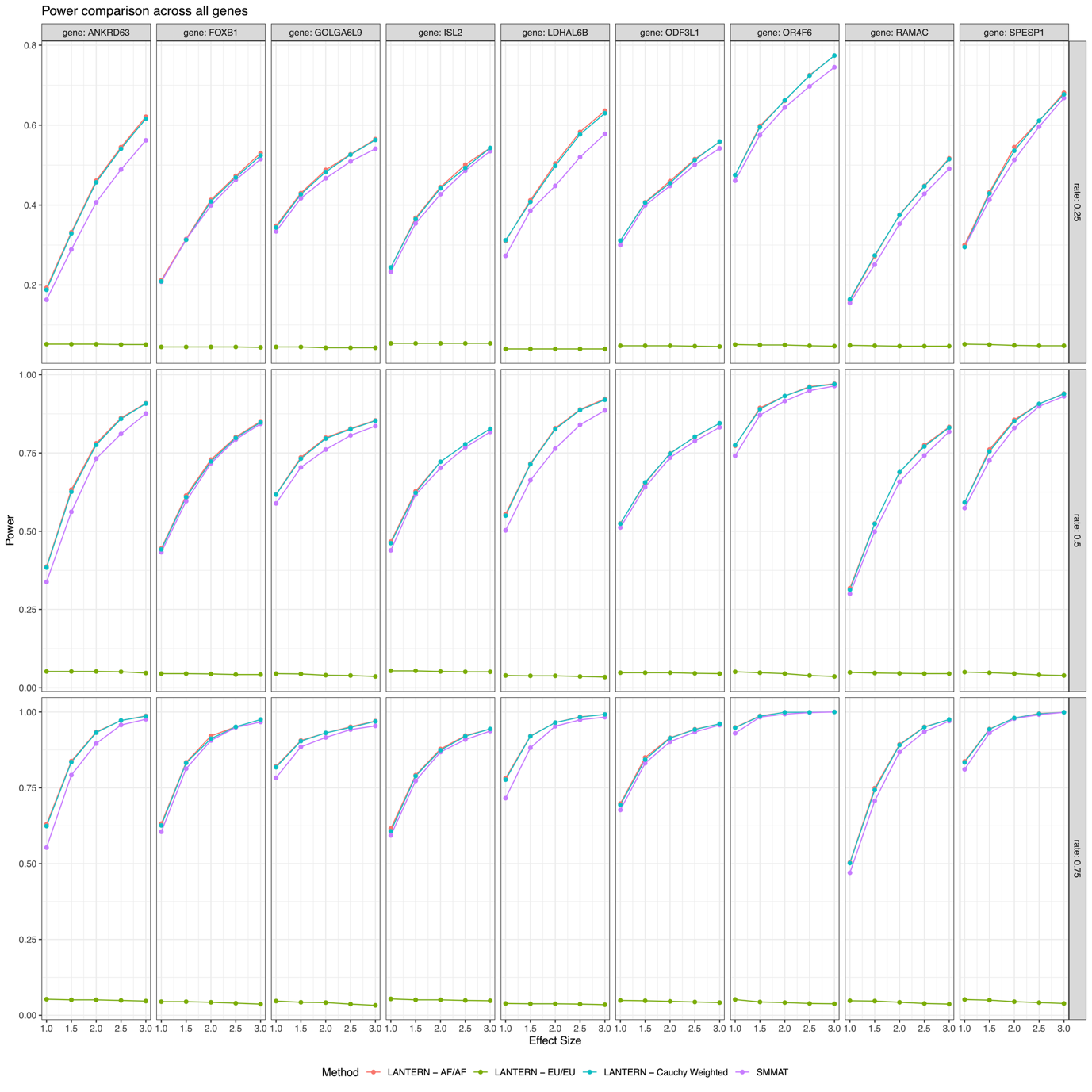

**Figure S5:** Power to detect rare-variant associations across all 9 genes. The results for LANTERN-AF are shown in red, LANTER-EU in green, LANTERN-Cauchy in blue, and SMMAT in purple. Causal variants were simulated in 25% of variants (top panel), 50% of variants (middle panel), and 75% of variants (bottom panel). Causal alleles were placed exclusively on AF haplotypes. Simulated effect sizes are shown on the horizontal axis.

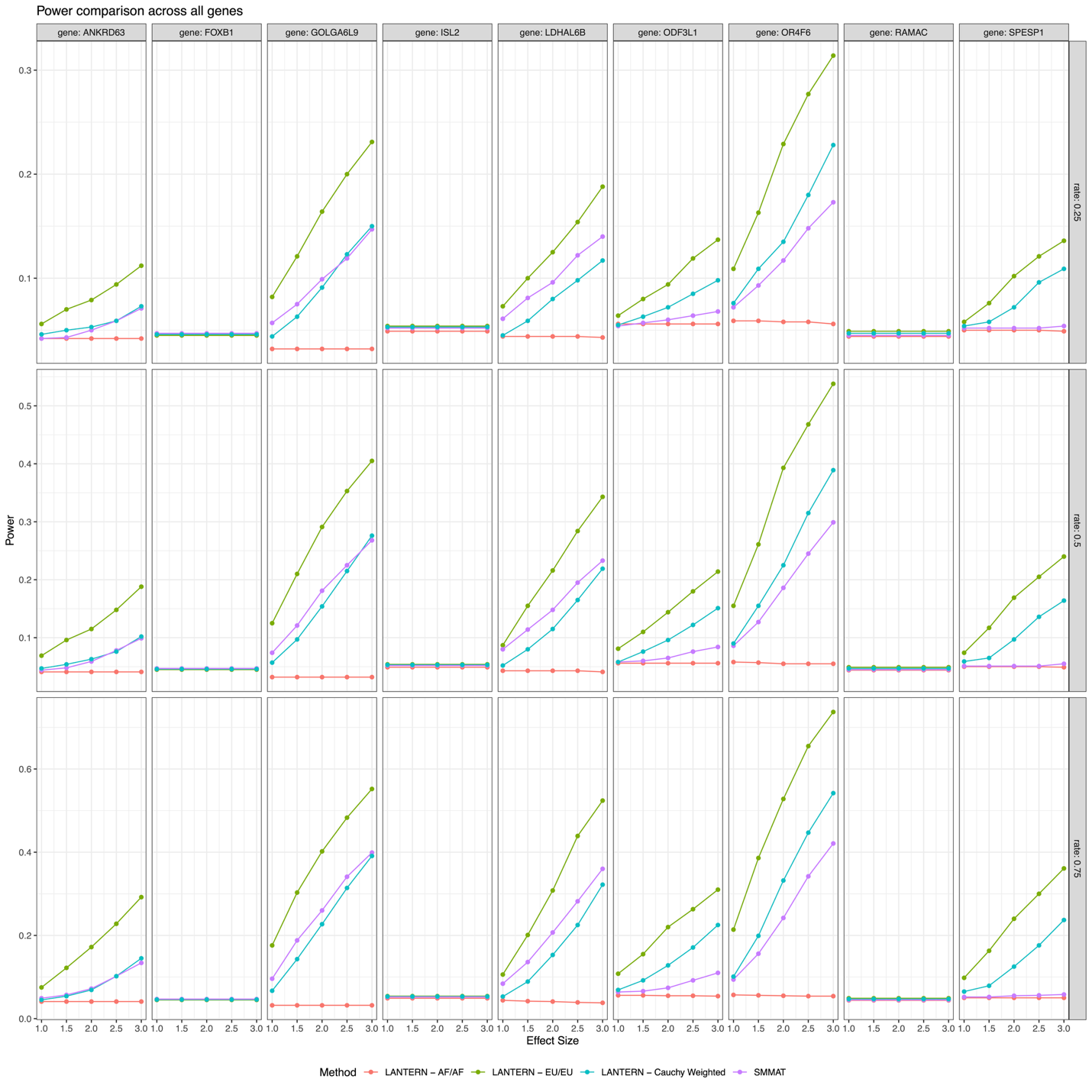

**Figure S6:** Power to detect rare-variant associations across all 9 genes. The results for LANTERN-AF are shown in red, LANTER-EU in green, LANTERN-Cauchy in blue, and SMMAT in purple. Causal variants were simulated in 25% of variants (top panel), 50% of variants (middle panel), and 75% of variants (bottom panel). Causal alleles were placed exclusively on EU haplotypes. Simulated effect sizes are shown on the horizontal axis.

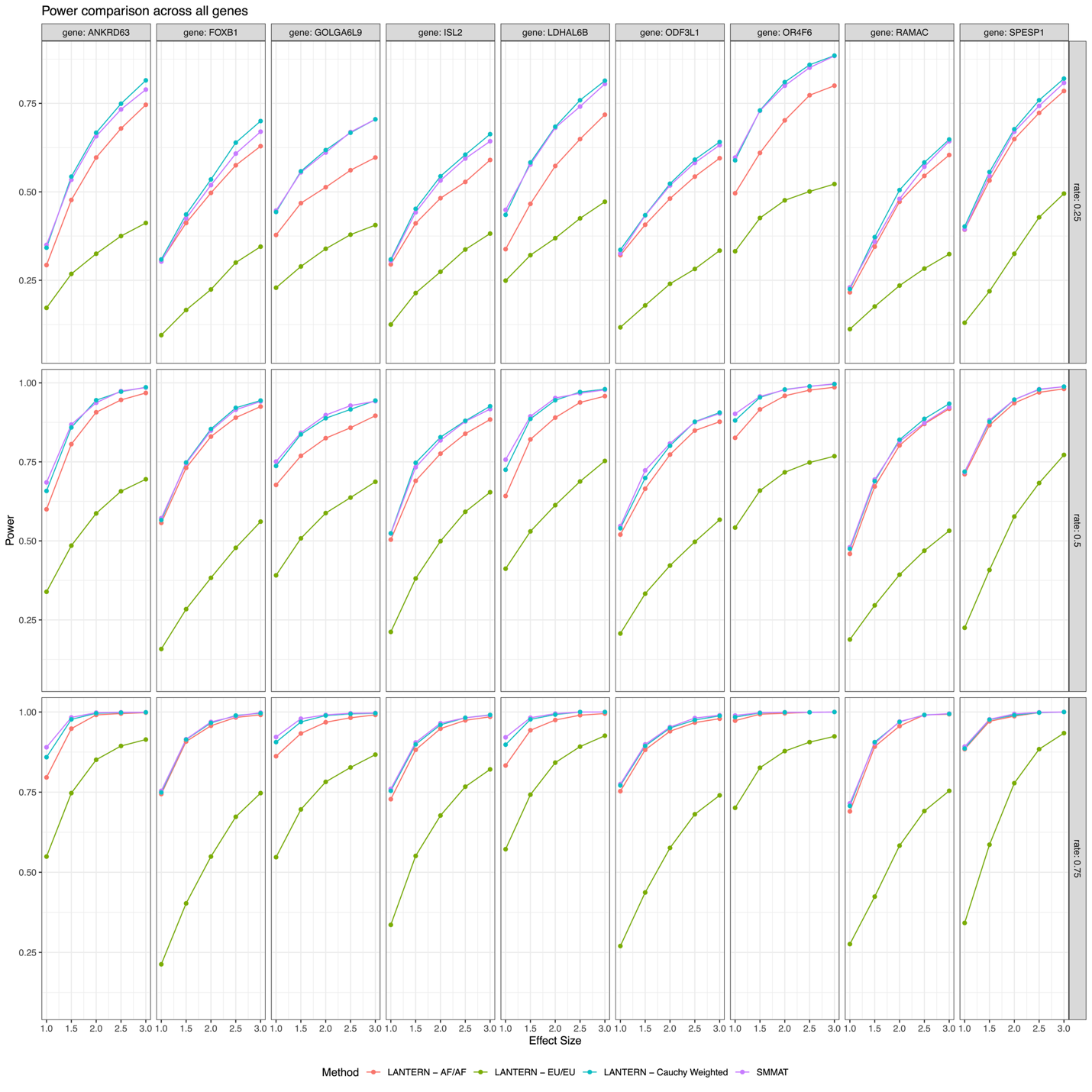

**Figure S7:** Power to detect rare-variant associations across all 9 genes. The results for LANTERN-AF are shown in red, LANTER-EU in green, LANTERN-Cauchy in blue, and SMMAT in purple. Causal variants were simulated in 25% of variants (top panel), 50% of variants (middle panel), and 75% of variants (bottom panel). Causal alleles simulated regardless of ancestral haplotype. Simulated effect sizes are shown on the horizontal axis.

| A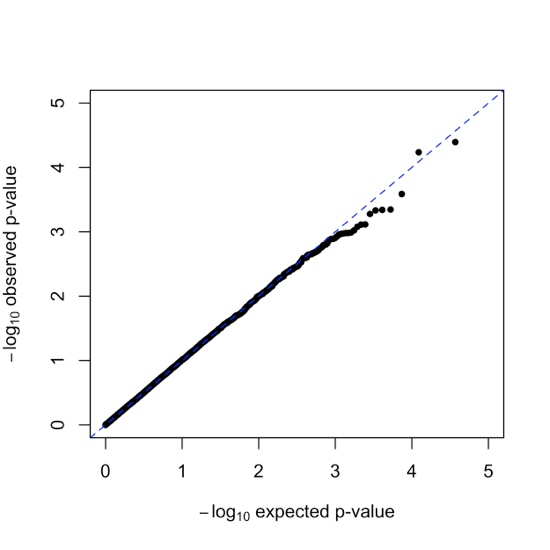 | B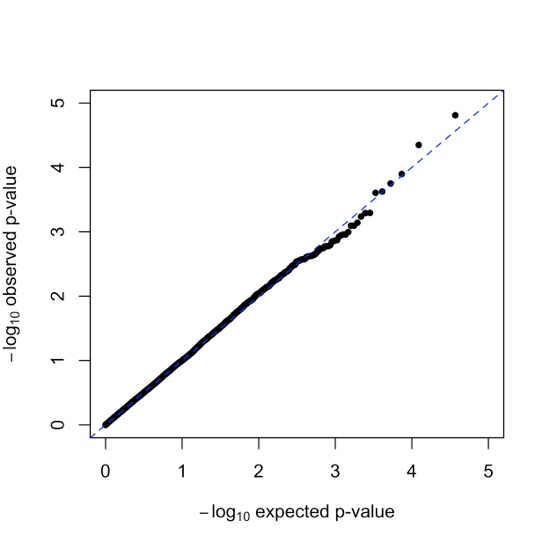 |
| --- | --- |
| C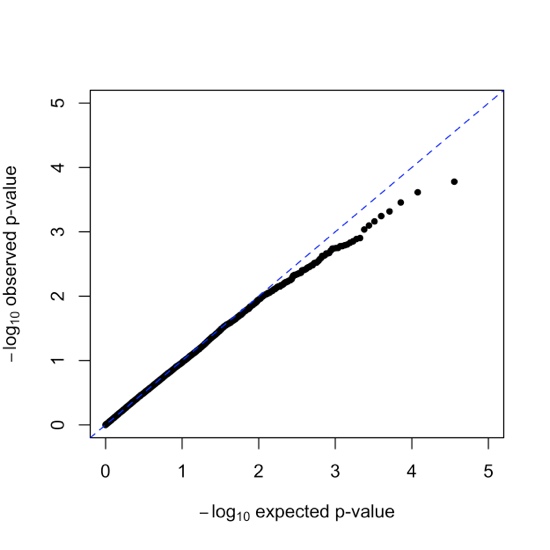 | D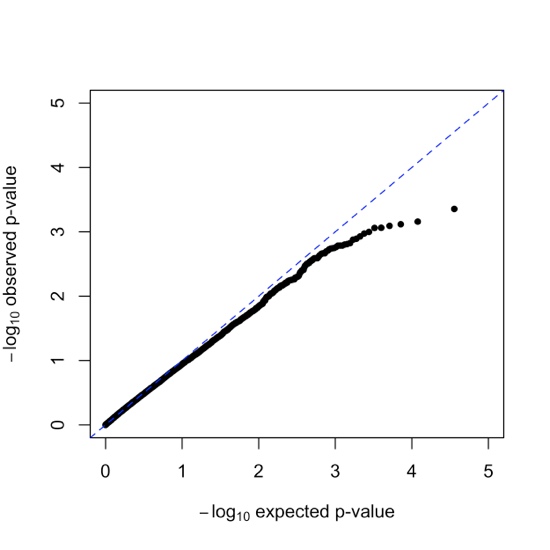 |
| E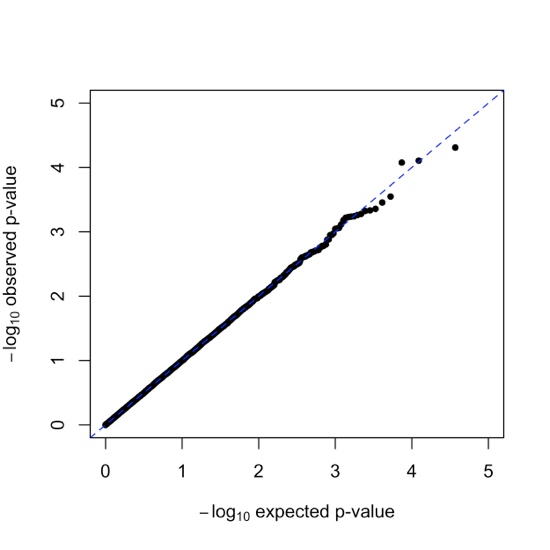 | 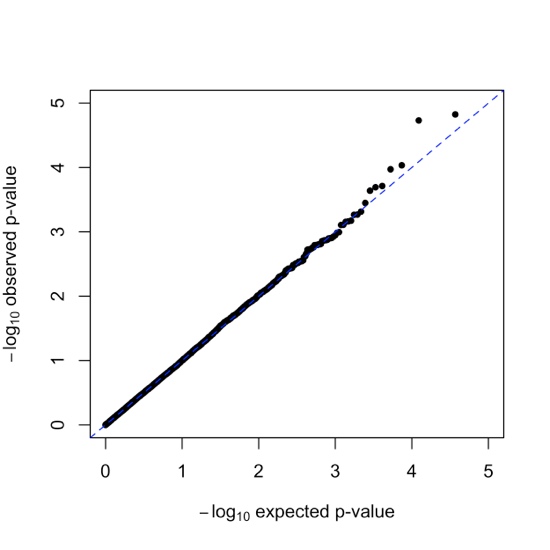F |

**Figure S8:** QQplots for exome-wide associations in JHS with HGB levels for the LANTERN-AF Burden test (A), LANTERN-AF SKAT-O test (B), LANTERN-EU Burden test (C), LANTERN-EU SKAT-O test (D), SMMAT Burden test (E), and SMMAT SKAT-O test (F).

| A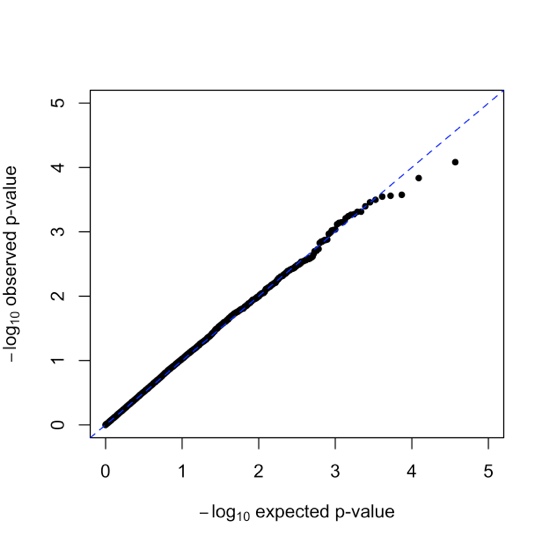 | 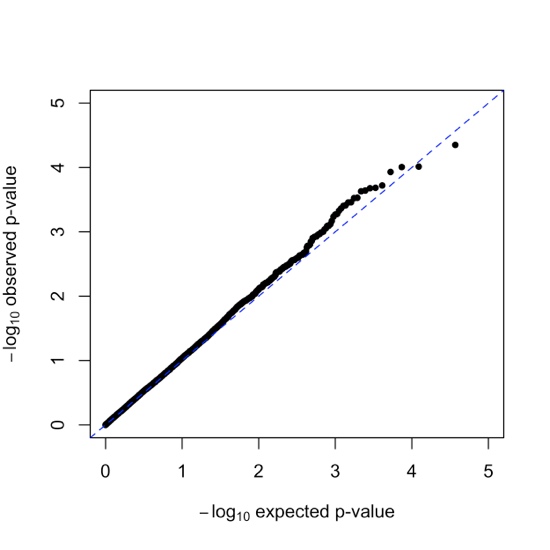B |
| --- | --- |
| C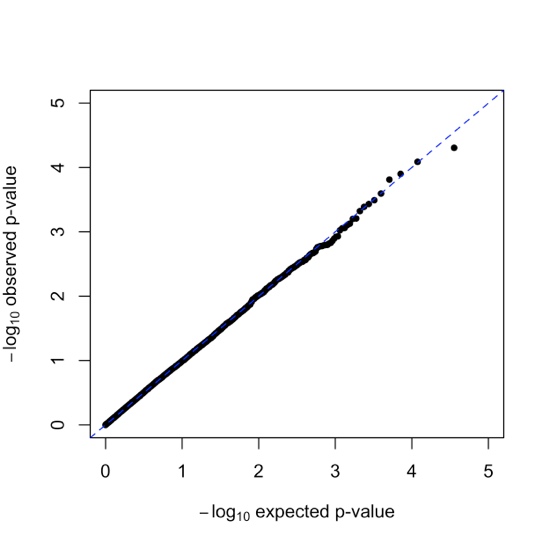 | D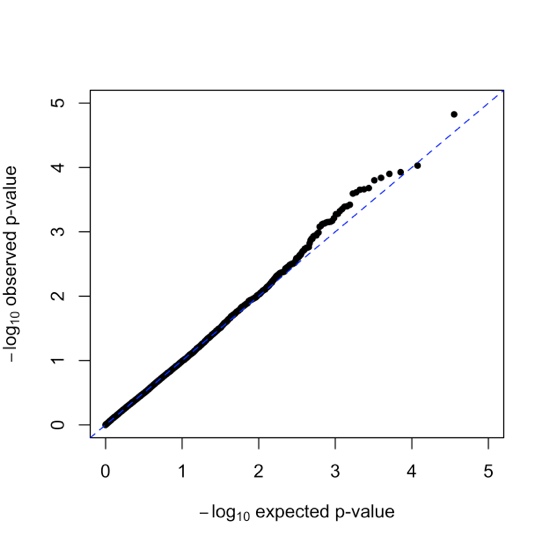 |
| E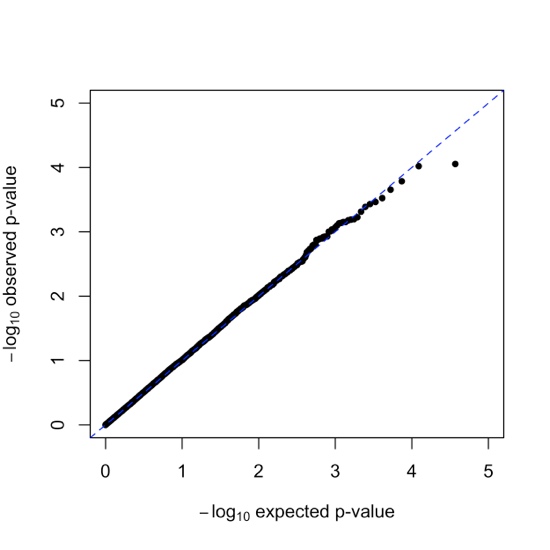 | F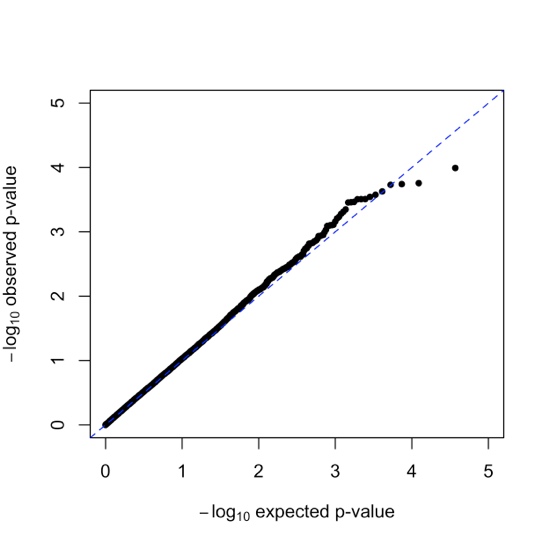 |

**Figure S9:** QQplots for exome-wide associations in JHS with RBC levels for the LANTERN-AF Burden test (A), LANTERN-AF SKAT-O test (B), LANTERN-EU Burden test (C), LANTERN-EU SKAT-O test (D), SMMAT Burden test (E), and SMMAT SKAT-O test (F).
